# Performance of reasoning large language models on nephrology multiple-choice questions

**DOI:** 10.64898/2025.12.03.25341427

**Authors:** Fumiya Kitano, Mamoru Masaki, Daisuke Ichikawa, Yugo Shibagaki, Ryunosuke Noda

## Abstract

**Aim:** Performance of large language models in medicine is improving, yet it remains unclear how the advantage of reasoning models depends on task characteristics in nephrology.

**Methods:** We evaluated four large language models in two families—OpenAI (GPT-5 reasoning, GPT-4o baseline) and Google (Gemini 2.5 Pro reasoning, Gemini 2.0 Flash baseline)—on 209 self-assessment questions for nephrology board renewal published by the Japanese society of nephrology. Questions were categorized by question type (general vs clinical), taxonomy (recall, interpretation, problem-solving), and image inclusion (non-image vs image). Models were assessed via application programming interface with default parameters; images were provided as PNG files. Accuracy used Wilson 95% confidence intervals (CIs); paired comparisons used McNemar’s exact test. Primary analyses used logistic generalized linear mixed models with fixed effects, random intercepts, and prespecified interactions.

**Results:** Overall accuracy was 87.6% (183/209, 95% CI 82.4–91.4) for GPT-5 and 83.7% (175/209, 95% CI 78.1-88.1) for Gemini 2.5 Pro vs. 69.9% (146/209, 95% CI 63.3-75.7) for GPT-4o and 62.7% (131/209, 95% CI 55.9-69.0) for Gemini 2.0 Flash. Paired analyses favored reasoning models, odds ratios of 6.29 for OpenAI and 7.29 for Google (both P<0.001). Adjusted odds ratios for reasoning vs. baseline were 5.00 for OpenAI and 7.28 for Google (both P<0.001). Interactions showed stronger effects in clinical questions for OpenAI and taxonomy-dependent effects for Google; no significant modification by image inclusion.

**Conclusion:** Reasoning models outperform baseline models with context-dependent advantages in nephrology, although their benefits vary by task and further validation is essential before routine use.

## 1. Introduction

Large language models (LLMs) have brought groundbreaking advances in natural language processing, demonstrating high performance in diverse tasks such as summarization, translation, question answering, and complex text generation [1,2]. In the medical field, they are expected to contribute widely to applications including medical education support, clinical decision-making, and research efficiency [3–5]. In particular, OpenAI’s GPT series and Google’s Gemini series have shown promising results in the United States Medical Licensing Examination, the Japanese National Medical Licensing Examination, and various specialty board examinations, demonstrating both medical knowledge and problem-solving ability [6–14].

Recently, reasoning models, designed to enhance stepwise logical thinking, have emerged [15–19]. These models incorporate techniques such as chain-of-thought prompting, self-consistency, external tool integration, and optimization for generating stepwise reasoning processes. They are specifically designed to maintain multi-step reasoning and logical consistency. In contrast, widely used earlier-generation LLMs (hereafter referred to as baseline models) primarily rely on large-scale text-based pattern learning, with noted limitations in explicitly presenting intermediate reasoning steps or supporting arguments [15]. Reasoning models can increase transparency by showing intermediate thought processes and facilitate error analysis, whereas they also present risks, such as increased computational costs and the tendency to produce plausible but incorrect reasoning (hallucinations) [20].

Prior studies have reported that baseline models such as GPT-4 may demonstrate certain capabilities in nephrology board-level questions, suggesting that LLMs could serve as useful tools for education and clinical practice support [13,14]. Nevertheless, nephrology is a highly specialized field that involves complex physiology related to systemic homeostasis, a broad disease spectrum, and the management of multiple comorbidities. In this domain, advanced clinical reasoning is indispensable, including the interpretation of test results, differential diagnosis based on pathophysiology, and formulation of individualized treatment strategies, all of which are suitable for reasoning models rather than baseline models. However, it has not been sufficiently investigated how the superiority of reasoning models is influenced by task characteristics in nephrology. Therefore, we aimed to quantify the performance gap between reasoning and baseline models in nephrology and to analyze how this gap is modified by specific question characteristics.

## 2. Methods

### 2.1. Self-Assessment Questions for Nephrology Board Renewal

The Self-Assessment Questions for Nephrology Board Renewal (SAQ-NBR) consist of nephrology-related multiple-choice questions written in Japanese, administered annually by the Japanese Society of Nephrology [21]. The SAQ-NBR is a required component for renewing nephrology board certification and is designed to comprehensively assess knowledge across the full spectrum of kidney diseases. The passing criterion is defined as an overall accuracy rate of ≥60% [22]. We analyzed 209 of 210 multiple-choice questions drawn from ten annual SAQ-NBR examinations administered between 2014 and 2023, after excluding one item that had been officially invalidated by the Japanese Society of Nephrology. Each question was annotated along four prespecified categories to capture task characteristics, defined in line with prior literature [13, 14]. Specifically, question type was classified as general (testing factual knowledge) or clinical (testing application in patient scenarios), and taxonomy (testing increasing levels of cognitive demand) was divided into recall, interpretation, and problem-solving (following the Japanese National Medical Examination taxonomy framework created by the Japan Medical Association [23]), image inclusion was defined as non-image vs. image-based questions (pathology micrographs or radiological images, and schematic diagrams), and subspecialty was mapped to eight domains (chronic kidney disease (CKD)/end-stage kidney disease (ESKD), acute kidney injury (AKI), glomerular diseases, tubulointerstitial diseases, hypertension/vascular diseases, water–electrolyte–acid–base disorders, autosomal dominant polycystic kidney disease (ADPKD)/urology, and basic medicine) using the Japan Society of Nephrology’s case experience list [24].

### 2.2. Large Language Models and Evaluation Settings

Two LLM families were evaluated, each comprising a reasoning model and a baseline model: OpenAI (GPT-5 (gpt-5-2025-04-16) as the reasoning model and GPT-4o (gpt-4o-2024-11-20) as the baseline) and Google (Gemini 2.5 Pro (gemini-2.5-pro-preview-03-25) as the reasoning model and Gemini 2.0 Flash (gemini-2.0-flash-001) as the baseline). To approximate production-like use, all models were accessed via application programming interface with default parameters and without temperature or top-p tuning. GPT-4o, Gemini 2.5 Pro, and Gemini 2.0 Flash were evaluated in April 2025, whereas GPT-5 was evaluated in August 2025. All questions were presented in Japanese, preceded by a standardized prompt: “We will now present a nephrology-related question. Please provide your answer and explanation.” For image questions, the corresponding PNG files were supplied alongside the text as multimodal inputs in the application programming interface call. For each model–question pair, the outcome was defined as a binary variable (correct = 1, incorrect = 0). Correctness was adjudicated against the official answer keys issued by the Japanese Society of Nephrology.

### 2.3. Statistical Analysis

We first computed overall and category-wise accuracy with Wilson 95% confidence intervals (CIs). Within each family, paired comparisons between the reasoning and baseline models were performed using McNemar’s exact test, and odds ratios (ORs) based on discordant pairs were reported. The primary analyses used logistic generalized linear mixed models (GLMMs) fitted separately by family. Fixed effects included model (reasoning vs. baseline), question type (general vs. clinical), taxonomy (recall, interpretation, problem-solving), and image inclusion (image vs. non-image). To assess effect modification, we prespecified interaction terms between model and question type, model and taxonomy, and model and image, with inference based on Wald tests. To accommodate the nested data structure, we included random intercepts for question and for case scenario in chained or related items. Because of concerns about multiplicity and estimator stability, subspecialty was not entered as an interaction term in the GLMMs. We reported adjusted odds ratios (aORs) with 95% CIs as the principal effect measures. For visualization, simple-effect aORs were derived from the GLMM coefficients and displayed in forest plots for the prespecified stratifications; All analyses were conducted in Python (version 3.11.13), and a two-sided p-value <0.05 was considered statistically significant.

## 3. Results

### 3.1. Overall performance and model comparisons

For the 209 questions analyzed, the overall accuracy of the reasoning models was 87.6% for GPT-5 (183/209, 95% CI 82.4–91.4) and 83.7% for Gemini 2.5 Pro (175/209, 95% CI 78.1–88.1). For the baseline models, GPT-4o achieved 69.9% (146/209, 95% CI 63.3–75.7) and Gemini 2.0 Flash 62.7% (131/209, 95% CI 55.9–69.0). Thus, in both the OpenAI and Google families, the reasoning models outperformed the baseline models (**Figure 1**). In paired comparisons on identical questions, GPT-5 was exclusively correct on 44 questions, while GPT-4o was exclusively correct on 7 (OR 6.29, 95% CI 3.25–16.00, p < 0.001). Similarly, Gemini 2.5 Pro was exclusively correct on 51 questions compared to 7 for Gemini 2.0 Flash (OR 7.29, 95% CI 3.83–18.33, p < 0.001). In both families, the reasoning models were significantly superior (**Table 1**).

**Figure 1.**
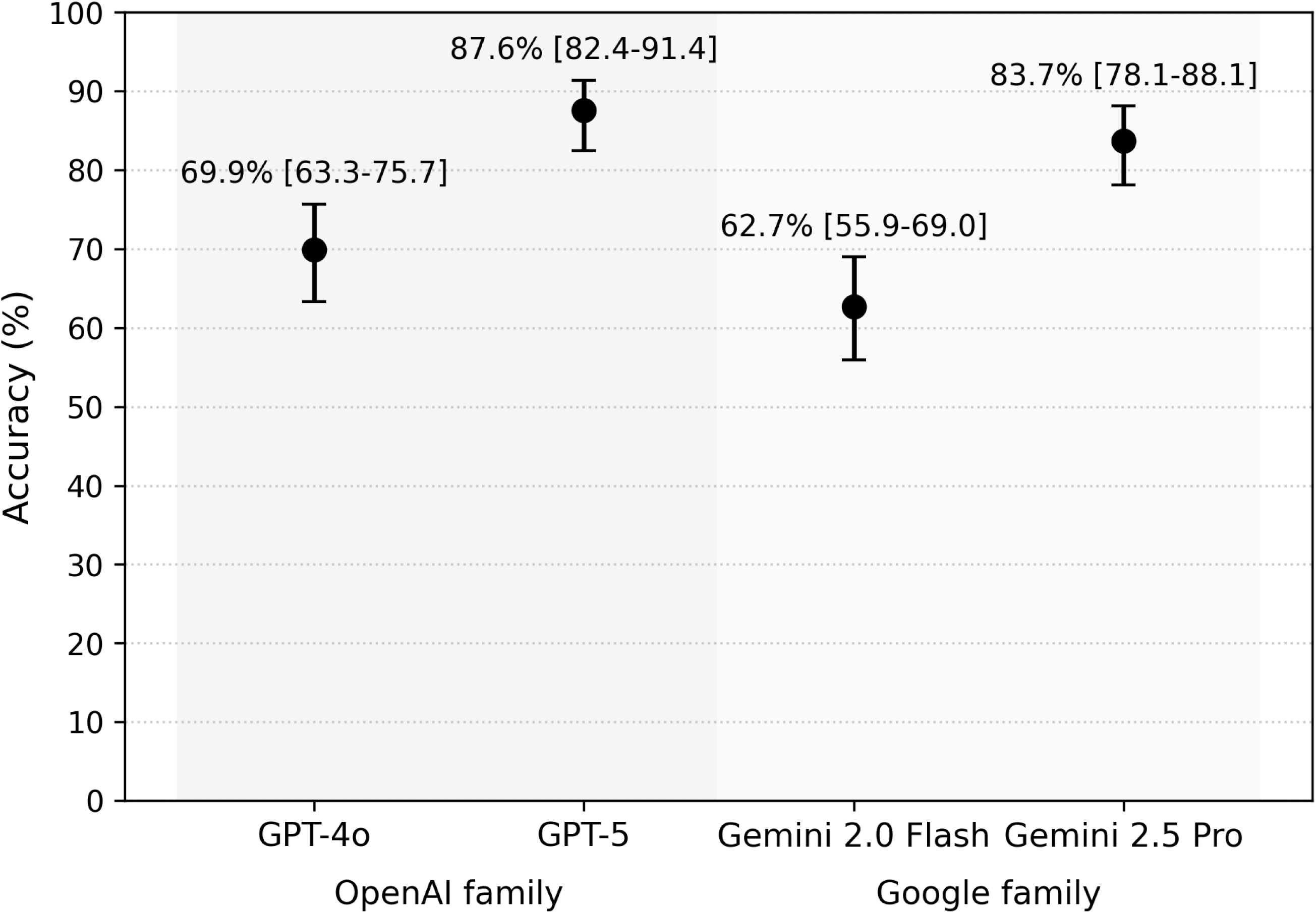
Overall accuracy on Self-Assessment Questions for Nephrology Board Renewal. For each model, the dot shows the percentage of correct answers and the error bars show the 95% Wilson confidence intervals. In both model families, the reasoning models (GPT-5 and Gemini 2.5 Pro) outperformed their respective baseline models (GPT-4o and Gemini 2.0 Flash).

**Table 1.** Paired comparison of performance between reasoning and baseline models.

| Comparison | Reasoning model<br>only correct (n) | Baseline model<br>only correct (n) | Risk difference | Odds ratio<br>(95% CI) | P-value |
| --- | --- | --- | --- | --- | --- |
| OpenAI (GPT-5 vs.<br>GPT-4o) | 44 | 7 | 0.18 | 6.29 [3.25-16.00] | < 0.001 |
| Google (Gemini 2.5<br>Pro vs. 2.0 Flash) | 51 | 7 | 0.21 | 7.29 [3.83-18.33] | < 0.001 |

The table displays a paired analysis comparing the performance of two model types on the same set of questions. The column "Reasoning model only correct (n)" indicates the number of questions answered correctly by the reasoning model but incorrectly by the baseline model. Conversely, "Baseline model only correct (n)" indicates the number of items answered correctly by the baseline model but incorrectly by the reasoning model. The Odds Ratio is calculated as the ratio of these two discordant pair counts. P-values were derived from McNemar's exact test. Abbreviations: CI, Confidence Interval.

### 3.2. Accuracy by question categories

When stratified by question categories, reasoning models consistently achieved higher accuracy across all major categories (Question Type, Taxonomy, and Image Inclusion) compared to baseline models. In the OpenAI family, the performance gap was particularly notable in clinical questions, where GPT-5 achieved 86.7% (95% CI 78.6-92.1) versus 65.3% (95% CI 55.5-74.0) for GPT-4o. In the Google family, a large gap was observed in recall questions, with Gemini 2.5 Pro at 86.1% (95% CI 78.3-91.4) compared to 62.0% (95% CI 52.6-70.6) for Gemini 2.0 Flash. Detailed stratified accuracies, including by subspecialty, are provided in **Table 2**.

**Table 2.** The proportion of the correct answers of GPT-4o, GPT-5, Gemini 2.0 Flash, and Gemini 2.5 Pro by four categories on the Self-Assessment Questions for Nephrology Board Renewal.

| Category | GPT-4o | GPT-5 | Gemini 2.0 Flash | Gemini 2.5 Pro |
| --- | --- | --- | --- | --- |
| *Question Type* |  |  |  |  |
| General | 73.9% [65.0-81.1]<br>(82/111) | 88.3% [81.0-93.0]<br>(98/111) | 64.0% [54.7-72.3]<br>(71/111) | 85.6% [77.9-90.9]<br>(95/111) |
| Clinical | 65.3% [55.5-74.0]<br>(64/98) | 86.7% [78.6-92.1]<br>(85/98) | 61.2% [51.3-70.3]<br>(60/98) | 81.6% [72.8-88.1]<br>(80/98) |
| *Taxonomy* |  |  |  |  |
| Recall | 73.1% [64.1-80.6]<br>(79/108) | 88.9% [81.6-93.5]<br>(96/108) | 62.0% [52.6-70.6]<br>(67/108) | 86.1% [78.3-91.4]<br>(93/108) |
| Interpretation | 66.1% [53.0-77.1]<br>(37/56) | 82.1% [70.2-90.0]<br>(46/56) | 60.7% [47.6-72.4]<br>(34/56) | 76.8% [64.2-85.9]<br>(43/56) |
| Problem-Solving | 66.7% [52.1-78.6]<br>(30/45) | 91.1% [79.3-96.5]<br>(41/45) | 66.7% [52.1-78.6]<br>(30/45) | 86.7% [73.8-93.7]<br>(39/45) |
| *Image Inclusion* |  |  |  |  |
| Non-Image | 71.6% [64.4-77.9]<br>(121/169) | 89.3% [83.8-93.2]<br>(151/169) | 61.5% [54.0-68.5]<br>(104/169) | 83.4% [77.1-88.3]<br>(141/169) |
| Image | 62.5% [47.0-75.8]<br>(25/40) | 80.0% [65.2-89.5]<br>(32/40) | 67.5% [52.0-79.9]<br>(27/40) | 85.0% [70.9-92.9]<br>(34/40) |
| *Subspecialty* |  |  |  |  |
| CKD/ESKD | 71.1% [56.6-82.3]<br>(32/45) | 84.4% [71.2-92.3]<br>(38/45) | 51.1% [37.0-65.0]<br>(23/45) | 80.0% [66.2-89.1]<br>(36/45) |
| AKI | 70.0% [39.7-89.2] (7/10) | 90.0% [59.6-98.2] (9/10) | 70.0% [39.7-89.2] (7/10) | 80.0% [49.0-94.3] (8/10) |
| Glomerular Diseases | 64.9% [51.9-76.0]<br>(37/57) | 86.0% [74.7-92.7]<br>(49/57) | 71.9% [59.2-81.9]<br>(41/57) | 89.5% [78.9-95.1]<br>(51/57) |
| Tubulointerstitial<br>Diseases | 63.2% [41.0-80.9]<br>(12/19) | 94.7% [75.4-99.1]<br>(18/19) | 47.4% [27.3-68.3] (9/19) | 89.5% [68.6-97.1]<br>(17/19) |
| Hypertension/Vascular<br>Diseases | 66.7% [43.7-83.7]<br>(12/18) | 72.2% [49.1-87.5]<br>(13/18) | 66.7% [43.7-83.7]<br>(12/18) | 72.2% [49.1-87.5]<br>(13/18) |
| Water/Electrolyte/Acid-Base Disorder | 60.9% [40.8-77.8]<br>(14/23) | 95.7% [79.0-99.2]<br>(22/23) | 65.2% [44.9-81.2]<br>(15/23) | 82.6% [62.9-93.0]<br>(19/23) |
| ADPKD/Urology | 91.7% [64.6-98.5]<br>(11/12) | 83.3% [55.2-95.3]<br>(10/12) | 75.0% [46.8-91.1] (9/12) | 75.0% [46.8-91.1] (9/12) |
| Basic Science | 84.0% [65.3-93.6]<br>(21/25) | 96.0% [80.5-99.3]<br>(24/25) | 60.0% [40.7-76.6]<br>(15/25) | 88.0% [70.0-95.8]<br>(22/25) |

Stratified accuracy by category (Wilson 95% CI). Values are percentage [95% CI] (correct/total). Abbreviations: CKD chronic kidney disease, ESKD end-stage kidney disease, AKI acute kidney injury, ADPKD autosomal dominant polycystic kidney disease.

### 3.3. Multivariable analysis adjusted for question categories

To assess whether the superiority of reasoning models was modified by question characteristics, we fitted multivariable logistic generalized linear mixed models (GLMMs) with prespecified interaction terms (**Table 3**). In the OpenAI family, we found a significant interaction between the model and Question Type (p for interaction = 0.007). The advantage of the reasoning model was significantly greater for clinical questions (aOR 13.94, 95% CI 5.68-34.25) compared to general questions (aOR 5.00, 95% CI 3.00-8.35) (**Figure 2a**).

**Figure 2.**
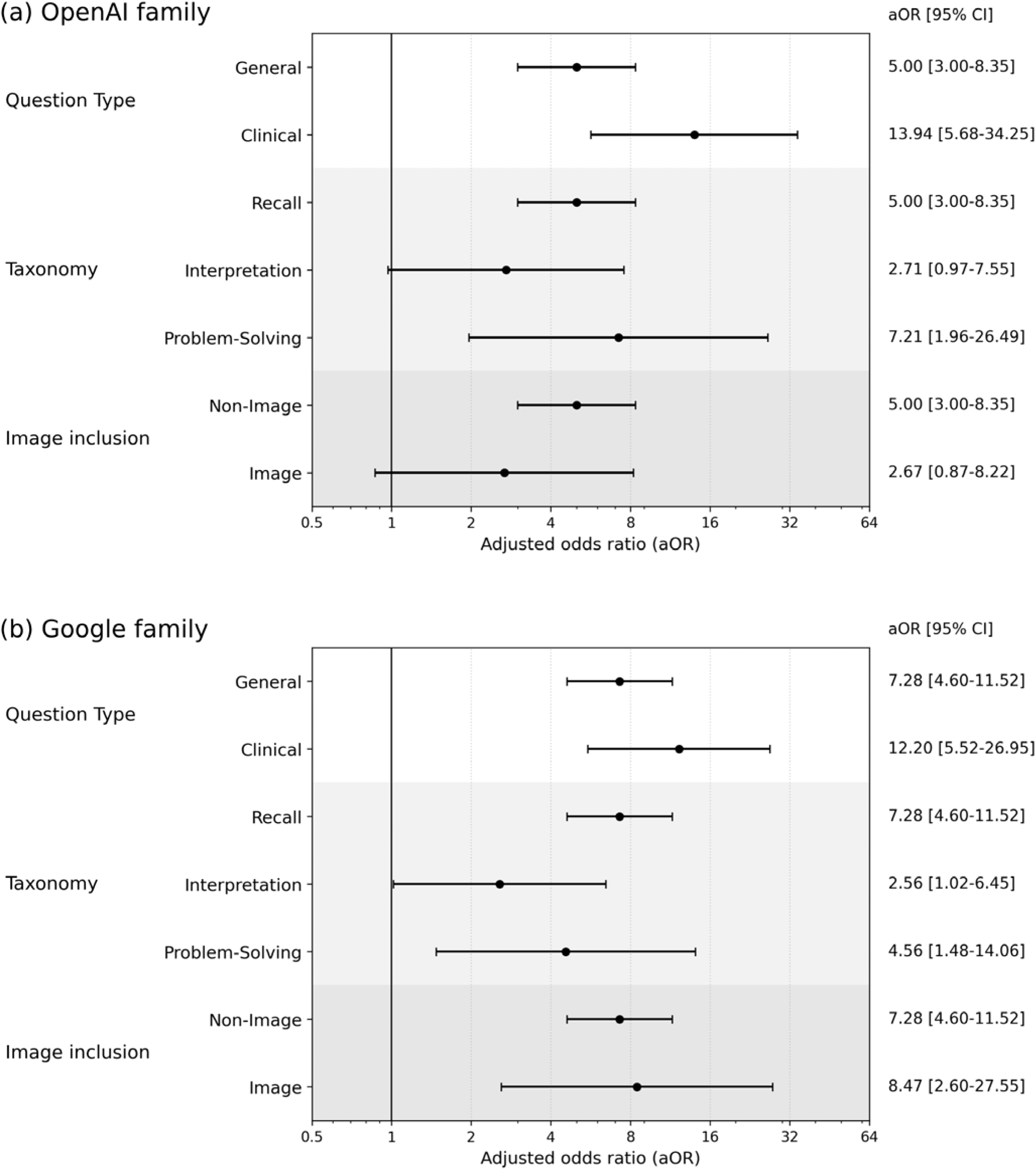
Subgroup effects of reasoning models vs. baseline models. Forest plots display adjusted odds ratios (aOR) for a correct answer by the reasoning models compared with the baseline models within (a) the OpenAI family and (b) the Google family. Estimates come from a logistic generalized linear mixed model adjusting for question characteristics (Question Type, Taxonomy, and Image Inclusion) with random intercepts for each question and case scenario. Subgroup values are simple effects relative to the reference category “General,” “Recall,” and “Non-Image.” An aOR >1 favors the reasoning model. Error bars indicate 95% confidence intervals and the x-axis is on a log scale.

**Table 3.**
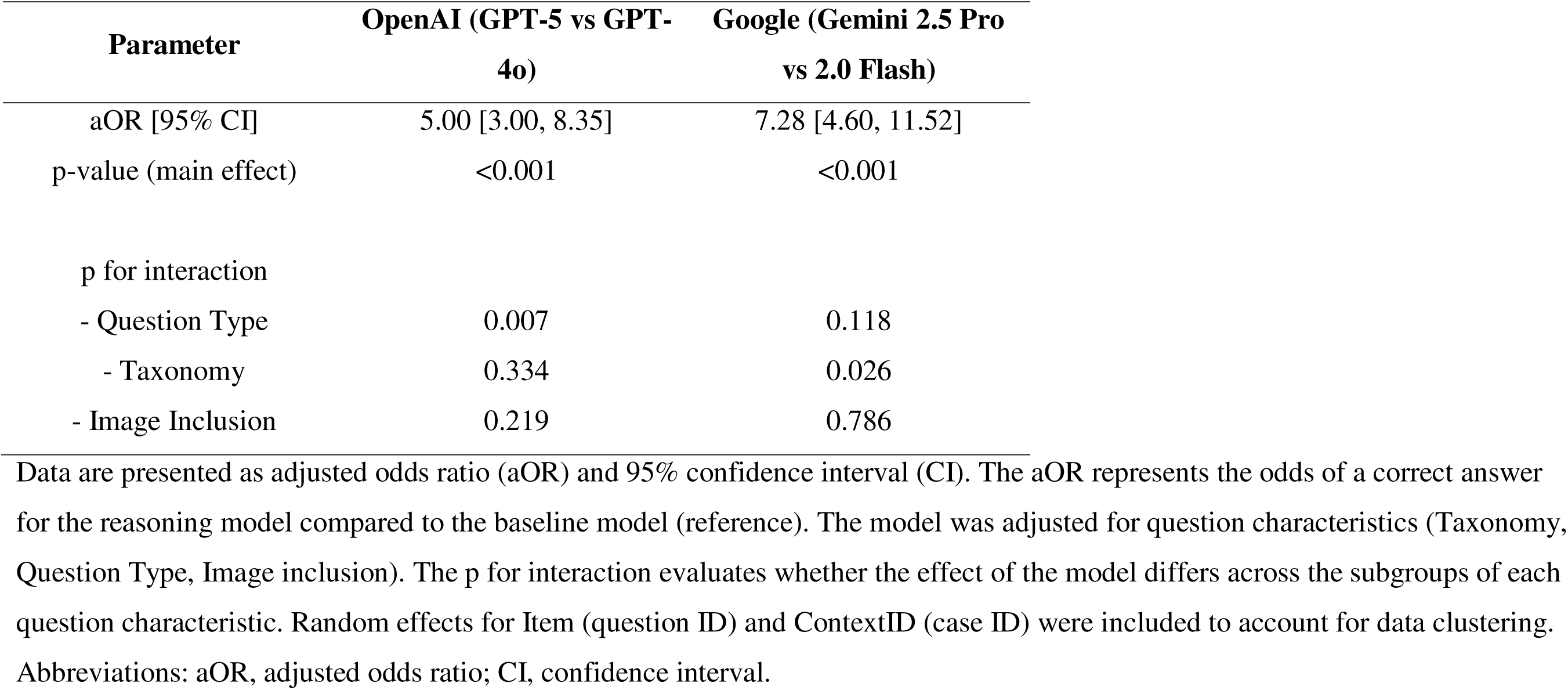
Comparison of large language model performance by multivariable generalized linear mixed model.

| **Parameter** | **OpenAI (GPT-5 vs GPT-4o)** | **Google (Gemini 2.5 Pro vs 2.0 Flash)** |
| --- | --- | --- |
| aOR [95% CI] | 5.00 [3.00, 8.35] | 7.28 [4.60, 11.52] |
| p-value (main effect) | <0.001 | <0.001 |
| p for interaction |  |  |
| - Question Type | 0.007 | 0.118 |
| - Taxonomy | 0.334 | 0.026 |
| - Image Inclusion | 0.219 | 0.786 |

Data are presented as adjusted odds ratio (aOR) and 95% confidence interval (CI). The aOR represents the odds of a correct answer for the reasoning model compared to the baseline model (reference). The model was adjusted for question characteristics (Taxonomy, Question Type, Image inclusion). The p for interaction evaluates whether the effect of the model differs across the subgroups of each question characteristic. Random effects for Item (question ID) and ContextID (case ID) were included to account for data clustering. Abbreviations: aOR, adjusted odds ratio; CI, confidence interval.

Conversely, in the Google family, a significant interaction was observed between the model and Taxonomy (p for interaction = 0.026). The reasoning model’s advantage was significantly smaller for interpretation questions (aOR 2.56, 95% CI 1.02-6.45) compared to the reference category of recall questions (aOR 7.28, 95% CI 4.60-11.52) (**Figure 2b**). Finally, no significant interaction was found with Image Inclusion in either the OpenAI family (p for interaction = 0.219) or the Google family (p for interaction = 0.786), indicating that the reasoning models did not provide a statistically significant additional benefit for questions containing medical images compared to text-only questions.

### 3.4. Qualitative analysis of incorrect response patterns

Of the 209 questions, 105 (50.2%) were answered correctly by all four models. Twenty-five questions (12.0%) were answered correctly by both reasoning models but incorrectly by both baseline models. In contrast, there were no questions (0%) where both baseline models answered correctly and both reasoning models failed. Fifteen questions (7.2%) were answered incorrectly by all four models. A qualitative review of these 15 questions revealed that they frequently involved knowledge specific to Japanese clinical guidelines or domestic epidemiological data (**Table 4**).

**Table 4.** Analysis of Representative Questions Incorrectly Answered by All Models.

| Question Summary | Error Category | Error of Models |
| --- | --- | --- |
| Risk stratification in the Japanese Society of Hypertension Guidelines for the Management of Hypertension 2019 | Japan-specific guidelines | The models identified "obesity" as a major risk factor based on international standards, failing to reflect the unique risk stratification of the JSH 2019 guidelines where obesity is not included in the primary/secondary risk tiers. |
| Epidemiology of anemia in patients with non-dialysis CKD in Japan | Japan-specific epidemiological data | The models judged based on general clinical trends (anemia worsens as CKD progresses), failing to reflect Japan-specific epidemiological data from a large-scale JSDT survey showing that about half of patients with G4/G5 CKD achieve the target Hb level. |
| Malignancies in dialysis patients in Japan | Japan-specific epidemiological data | The models responded based on general cancer epidemiology, failing to reflect the unique epidemiology from Japanese registry data, which shows that genitourinary malignancies are the most common cancer in male dialysis patients. |
| Population attributable risk of cardiovascular mortality in the EPOCH-JAPAN study | Japan-specific epidemiological data | While acknowledging the importance of blood pressure control, the models failed to provide the specific population attributable risk (~40%) reported by the Japanese EPOCH-JAPAN study. |

The table presents a qualitative analysis of questions that all four language models answered incorrectly. For each representative problem, it provides a summary of the topic, classifies the error category, and describes the presumed reasoning for the models'
failure. The errors frequently stemmed from a reliance on international or generalized medical knowledge, which was inconsistent with clinical guidelines or epidemiological data specific to Japan.

## 4. Discussion

In this study using Japanese nephrology board renewal questions, reasoning models (GPT-5 and Gemini 2.5 Pro) consistently outperformed baseline models (GPT-4o and Gemini 2.0 Flash) in both the OpenAI and Google families. Their advantage was not consistent across all question types: in multivariable analyses, the magnitude of this advantage varied by question characteristics, with the OpenAI family showing particularly strong gains on clinical questions and the Google family showing notable improvements on recall questions. Questions testing Japan-specific guidelines or epidemiology were frequently answered incorrectly by all models, underscoring important challenges that must be addressed before these systems can be reliably applied in real-world clinical practice.

The overall superiority of reasoning models in this study is consistent with prior evaluations in which reasoning models achieved higher answer accuracy. For example, o1 pro achieved 81.3% answer accuracy compared with 51.2% for GPT-4 on nephrology board renewal multiple-choice questions [14], and o3 achieved 83.3% accuracy compared with 70.0% for ChatGPT-3.5 on an original set of pharmacotherapy MCQs [25]. In our study, the advantage of reasoning models was greater for clinical questions than for general questions, especially within the OpenAI family. This pattern suggests that these models may be particularly effective in tasks that require integrating multiple pieces of patient information and test data in context to approximate clinical decision-making. A similar possibility has been highlighted in an editorial discussing o1-preview, which suggests that its enhanced chain-of-thought reasoning could be beneficial for complex clinical case–based diagnostic and management tasks compared with GPT-4 [26].

Despite these overall performance gains, the benefits of reasoning models were not uniform across all task types, highlighting important limitations of current systems. In both the OpenAI and Google families, the advantages of reasoning models were more pronounced for basic recall questions but diminished for higher-order interpretation questions. Reasoning models can exhibit hallucinations, which can be accompanied by breakdowns in the reasoning process [20]. These models optimize for next-token fluency rather than verification of correctness, and longer chain-of-thought reasoning may allow small intermediate errors to accumulate into globally incorrect but superficially plausible explanations. This mechanism may partly explain why their advantages were diminished on interpretation questions. For baseline LLMs, prior work has shown generally favorable performance overall, but also notable errors on clinically complex, context-dependent questions [14]. One diagnostic accuracy study found that models such as GPT-4 achieved substantially lower scores on reasoning-type questions than on knowledge-based questions [27], and a separate investigation of clinical decision-making showed that non-reasoning LLMs displayed notable limitations in multi-step, real-world-like scenarios even when they achieved high scores on medical licensing-style examinations [28].

Our analysis revealed no significant interaction between reasoning model and image inclusion, even though image questions were expected to rely more on higher-level visual–textual reasoning. One possible explanation is that the integration of visual information may still be suboptimal. Even in the latest reasoning models such as GPT-5 and o3, recent studies have reported that performance in image-based spot-diagnosis tasks remains far below that of human specialists [29]. Similarly, evaluations using the NEJM Image Challenge and various specialty board examinations have shown that adding image inputs does not necessarily improve performance, and in some instances may offer no advantage over text-only inputs [30]. Other studies have further suggested that text-only descriptions without accompanying image can yield comparable or even superior accuracy [31, 32]. The true multimodal capabilities of reasoning LLMs that integrate image and text inputs remain unclear [33, 34].

Our findings suggest that current systems may still face important challenges in interpreting complex nephrology cases and in integrating visual and textual information for clinical reasoning.

Despite overall high accuracy, some questions were missed by all models, often involving Japan-specific guidelines or epidemiological data. These findings likely reflect insufficient alignment between LLMs and locally adapted guidance that depends on Japan-specific clinical information. This underscores a key limitation of LLMs trained primarily on English-language or broadly international data [35, 36]. Consistent with these findings, a systematic review of 45 studies across 17 countries reported that GPT-4 achieved high overall accuracy but performed significantly better on examinations from English-speaking countries, and that translating questions into English improved ChatGPT-3.5 but had no effect on GPT-4, underscoring persistent language- and context-dependent variability in performance [37].

Improving reliability will require mechanisms that reference local guidelines, cite sources, and indicate uncertainty. Retrieval-Augmented Generation could help address this, but quality control and verification are essential to ensure clinical safety [38, 39].

Several limitations were identified in the design and evaluation. First, the evaluation was limited to MCQs, constraining generalizability to open-ended, multi-turn clinical tasks [4]. MCQ-based designs can sometimes be solved through pattern recognition rather than genuine clinical reasoning. Prior reports using alternative assessment formats suggest that current LLMs may struggle when information is incomplete, conflicting, or clinically subtle, underscoring the need to evaluate not only whether they choose the right answer, but how they interpret and weigh clinical clues [40, 41]. Second, the SAQ-NBR dataset is publicly available, so data leakage into training corpora cannot be excluded [42]. Third, the scope was restricted to Japanese nephrology questions and within-family comparisons, limiting external validity. Finally, reproducibility is challenged by stochastic decoding and rapid model iteration; our findings reflect model snapshots from April to August 2025. These limitations indicate that future work should test LLMs in more realistic clinical scenarios, such as narrative cases and tasks requiring management decisions. Such evaluations will help clarify how these models can safely support nephrologists beyond simple factual recall.

In conclusion, reasoning models consistently outperformed baseline models on nephrology board-style questions, with strengths that varied by task characteristics. They may serve as useful adjuncts for education and decision support, but careful, task-specific validation will be essential before routine use. These findings clarify both the capabilities and limitations of LLMs in nephrology and support their responsible development and application in clinical practice.

## Research involving Human Participants and/or Animals

This article does not contain any studies with human participants or animals performed by any of the authors.

## Informed consent

Not applicable.

## Use of AI in manuscript preparation

Portions of the manuscript (translation and language editing) were assisted by an AI tool. All authors reviewed and take full responsibility for the content.

## Acknowledgements

We thank the Japanese Society of Nephrology for making SAQ-NBR question sets publicly available online, and colleagues who provided feedback on study design and analysis.

## Data availability and transparency

Because SAQ-NBR materials are proprietary, the raw questions and official answers cannot be shared. The complete analysis code that supports the findings of this study is available from the corresponding author upon reasonable request.

